# Evaluating the use of hierarchical composite endpoints in pediatric cancer supportive care clinical trials: Illustrative examples from two multi-center phase-III randomized clinical trials

**DOI:** 10.64898/2026.01.20.26344064

**Authors:** Willem H. Collier, Mark Zobeck, Adam J. Esbenshade, Christopher C. Dvorak, Lillian Sung, David Freyer, Sarah Alexander, Etan Orgel, Nicole J. Ullrich, Zach Prudowsky, Brian Fisher, Caitlin W. Elgarten

## Abstract

**PURPOSE:** Pediatric supportive care trials frequently rely on analyses of multiple clinically relevant outcomes, posing challenges for overall trial interpretation. Hierarchical composite endpoints (HCEs) rank relevant outcomes by prespecified clinical importance and offer potential advantages such as harmonizing trial conclusions.

**METHODS:** We reanalyzed two randomized supportive care trials utilizing post-hoc HCE, each conducted through the Children’s Oncology Group. ACCL0934 evaluated levofloxacin for prevention of bloodstream infection (BSI) in patients with acute leukemia (AL) or undergoing hematopoietic cell transplant (HCT) and was chosen because of observed multidimensional benefits of levofloxacin. ACCL0431 evaluated sodium thiosulfate (STS) for prevention of cisplatin-induced hearing loss. ACCL0431 analyses were performed for overall and localized disease subgroups, chosen due to conflicting effect directionality on hearing vs survival by cohort. We estimated treatment effects on HCEs using win-odds ratios (WO). For ACCL0934, the primary HCE included death, severe infection, BSI, and neutropenic fever. For ACCL0431, the HCE included death, relapse/progression, and hearing loss.

**RESULTS:** In ACCL0934, levofloxacin reduced BSI incidence, but only with corresponding p-value <0.05 in the AL cohort (22% vs 43% on control; *P*=0.003; HCT: 11% versus 17% on control; *P=*0.06). Using HCE reanalysis, the estimated win-odds achieved statistical significance in both cohorts (AL: WO=1.74, *P=*0.002; HCT: WO=1.28, *P=*0.031). In ACCL0431, HCE analyses resulted in an estimated null effect (WO=1) in the overall cohort but resulted in beneficial effects (WO>1) for analyses of the localized cohort.

**CONCLUSION:** HCEs can provide a harmonized framework for interpreting complex supportive care trials by integrating outcomes of varying clinical importance. These post-hoc analyses should not be used to reinterpret either trial but motivate consideration of prospective use of HCE going forward.

## Introduction

Optimizing outcomes for pediatric cancer patients relies on the successful advancement of evidence-based supportive care to ease treatment burden from toxicity and mitigate the long-term side-effects of cancer therapy. The Children’s Oncology Group (COG) Cancer Control (CCL) Committee oversees the design and completion of pediatric supportive care studies, primarily randomized clinical trials.^1^ Interpreting supportive care trials remains challenging, at least in part, due to a multiplicity of key clinical outcomes identified *a priori* as integral to interpretation of intervention effect.^2–4^ For example, a supportive care trial may evaluate a bacterial infection prophylaxis strategy, where hypothesized benefits include reductions in neutropenic fever episodes, empiric antibiotic exposure, bacteremia, and/or death. Conversely, hypothesized harms of a prophylaxis strategy may include increased incidence of *Clostridioides difficile* infection and antibacterial resistance. Moreover, overall disease outcomes (e.g., relapse free survival) may supersede the importance of supportive care outcomes (e.g., reduced infection or other toxicity), which can necessitate assessment of whether an intervention interferes with cancer-directed treatment efficacy. In this paper, we discuss how this outcome plurality can uniquely complicate interpretation of pediatric supportive care trials, and we overview and evaluate an analytical approach which can be used to address these challenges.

Traditionally, separate analyses are performed for each distinct outcome identified as relevant to evaluate comparative effectiveness in a supportive care clinical trial.^2–4^ This approach leads to relatively straightforward implementation and interpretation of each individual analysis. However, pediatric supportive care trials are often only powered for analysis of an *a priori* defined primary outcome, often leading to underpowered analyses of other pertinent outcomes.^3,5^ Underpowered analyses are especially common when conducting trials for rare diseases, such as in pediatric cancer. In addition, use of separate analyses on distinct outcomes can lead to the possibility of conflicting findings of benefit and harm of randomized intervention. Another approach to handling outcome multiplicity is the use of a traditional composite endpoint. This approach can be problematic as these endpoints typically impart clinical equivalence to each component. Such clinical equivalence may not be accurate, and an analysis that uses such a composite can mask the treatment effect on an outcome that is clinically important but occurs less frequently than others. The hierarchical composite endpoint (HCE) is an endpoint construction strategy developed to overcome such challenges.^6^ HCEs are a composite wherein each outcome is *a priori* ranked according to clinical importance. In the analysis, patients are compared using this hierarchy, which effectively gives higher weight to the more clinically important outcomes in determining the treatment effect size. Initially devised for trials of heart failure to analyze heart failure-related hospitalization and time-to-cardiovascular death in a single primary endpoint,^6,7^ HCEs have gained traction and have subsequently been used in pivotal phase III trials, including, in some cases, in support of regulatory approvals.^7–13^

We hypothesize that HCE can be applied to pediatric supportive care trials to improve the interpretability of the overall analysis of multiple outcomes. To explore this, we leveraged data from two previously completed and analyzed trials conducted under the purview of the COG CCL committee: ACCL0934, a trial assessing the efficacy of an antibacterial prophylaxis strategy and ACCL0431, a trial assessing the efficacy of an agent intended to prevent ototoxicity.^2,3^ The trials were chosen because of key illustrative differences. ACCL0934 has a short, well-defined follow-up period and an HCE can be constructed to represent a natural hierarchy of infection-related outcomes.^2^ ACCL0431 was chosen to evaluate the use of HCEs in a trial which demonstrated efficacy of investigational treatment on the primary supportive care outcome, but balanced by a higher incidence of adverse outcomes observed among certain patients randomized to treatment.^3^ To evaluate the use of this methodology, we developed theoretical, post-hoc HCEs and reanalyzed trial data using those HCEs under win statistical analysis. The results of these analyses provide an initial empirical assessment of HCE to inform trial design and statistical methodology for future HCE-amenable supportive care trials.

## Methods

### Overview of Approach to HCE Construction

Constructing an HCE is a two-step process. First, the key outcomes of the trial, to be used in interpreting the comparative effectiveness of two treatment options under consideration, are defined as HCE components. Second, those components of the HCE are ranked in order of clinical importance. It is imperative that each individual HCE component be clinically important should any one component strongly drive the treatment effect size observed on the overall HCE. Outcomes should be ranked based on clinical judgement and patient-centered priorities with consensus opinion from multiple stakeholders (e.g., patient advocates, clinicians, regulatory agencies, etc.). HCE construction is context specific, with unique considerations for each trial setting. Researchers can consider further guidance in the existing literature (e.g., HCEs in cardiology^7^, in infectious disease related contexts^13,14^, stem cell transplant^8^, and considerations for censoring and competing events^15^).

### Overview of Win-Statistic Analyses

To quantify the overall effect of randomized treatment assignment on the HCE chosen, we utilized a class of metrics, termed “win statistics,” developed specifically to quantify randomized treatment effects on HCE.^6,7^ Win-statistics are flexible in that they can handle HCE components of any data type (e.g., time-to-event, binary, continuous). In certain settings, win-statistics can be inferred upon using non-parametric methods (limiting mathematical assumptions).^6,15–18^ Among several frequently used win-statistic metrics in the literature such as the win-ratio, net benefit, and win-odds ratio, we selected the win-odds ratio due to its interpretability in the presence of “ties” (defined below).^19,20^ Win-odds ratios reflect the odds that a randomly selected patient from the intervention arm has a better overall outcome based on the HCE than a randomly selected control arm patient, and a win-odds greater than 1 implies benefit of randomization to the active relative to control option.^20^

We briefly illustrate the non-parametric approach to estimation of win-statistics, as used for this paper. Firstly, define P_1_ as the probability a randomly selected treatment arm patient has a better overall outcome, defined by the HCE, than a randomly selected control arm patient; P_2_: The probability a randomly selected control arm patient has a better overall outcome than a randomly selected active arm patient; P_tie_: The probability of a tie. The win-odds ratio is (P_1_+ P_tie_)/(P_2_+ P_tie_). For a fixed follow-up period for which every patient has non-missing data on HCE components, these quantities can and for this paper were estimated by performing a head-to-head comparison of each possible pair of active and control arm patients. For, example, for P_1_, we calculated the proportion, across all possible pairwise comparisons of treatment and control arm patients, for which the treatment arm patient “wins” over the control arm patient based on the following algorithm: Each two patients were first compared using the most clinically important HCE component and if a win (e.g., the treatment arm patient does not experience the most severe/clinically important HCE event but the control arm patient does) cannot be determined, the patients were compared with respect to the second component, and so on. If, across all HCE components, a winner could not be determined, the head-to-head comparison was a tie. P_2_ and P_tie_ were estimated under the same algorithm.

### Summary of Included Randomized Trials

Both ACCL0934 and ACCL0431 have been previously described, and additional detail is provided in the supplemental materials.^2,3^ In summary, ACCL0934 was a multi-center, open label, randomized phase 3 trial comparing levofloxacin versus no prophylaxis (with 1:1 randomization) for prevention of bloodstream infections (BSI) in patients undergoing at least one cycle of chemotherapy (excluding conditioning for transplant) for treatment of acute myeloid leukemia or relapsed acute lymphoblastic leukemia (Cohort 1), and separately for those undergoing hematopoietic cell transplantation (HCT, Cohort 2). The primary outcome of ACCL0934 was centrally adjudicated BSI, with several additional secondary, exploratory, and post-hoc outcomes collected, including neutropenic fever (FN, incidence and duration), severe infection, additional severe adverse events (e.g., CTCAE grade 4), *Clostridioides difficile*–associated infection, and death. Patients were followed during what was termed the infection observation period, approximately tied to the period of severe neutropenia (ANC<200 cells/μL) following relevant cancer directed treatment initiation.

ACCL0431 was a multi-center, open label randomized phase 3 trial of sodium thiosulfate (STS) for the prevention of cisplatin-induced hearing loss. Patients with different malignancies intended for treatment with cisplatin were enrolled (e.g., neuroblastoma, medulloblastoma, osteosarcoma, etc.). As such, the trial enrolled a heterogeneous group of patients, for example those with localized or disseminated disease at baseline. Participants were randomized 1:1 to STS or control (no STS). Trial endpoints included hearing loss following therapy, renal injury, hematologic toxicities, cancer relapse, progression, secondary malignancy (SMN), and death. Patients were followed for the ototoxicity primary outcome up-to approximately four weeks following completion of cisplatin therapy, and for death, relapse, progression, and secondary malignancies, until the trial was administratively completed.^3^

### Data Analyses Performed

For the reanalysis of both trials, we constructed two HCEs (Table 1). Additional rationale for HCE construction along with outcome definitions are provided in supplemental materials.

**Table 1:** Endpoint Summary.

| **Endpoint Construct** | **ACCL0934** | **ACCL0431** |
| --- | --- | --- |
| <u>Original Primary Endpoint</u> | 1. True bacteremia (binary endpoint) | 1. SIOP Grade $\geq 2$ Hearing Loss (binary endpoint) |
| <u>HCE Extension 1:</u> | Patients compared according to the following hierarchy, with decreasing level of clinical significance:<br>1. All cause death (binary)<br>2. Severe infection* (binary)<br>3. True bacteremia (binary)<br>4. Neutropenic fever days (continuous)<br>*Bacterial and non-bacterial | Patients compared according to the following hierarchy, with decreasing level of clinical significance:<br>1. Death+ (time-to-event)<br>2. Cancer relapse, progression, or SMN (time-to-event)<br>3. SIOP grade $\geq 2$ hearing loss (time-to-event)<br>+All-cause |
| <u>HCE Extension 2:</u> | Patients compared according to the following hierarchy, with decreasing level of clinical significance:<br>1. All cause death (binary)<br>2. Severe infection* (binary)<br>3. Other serious adverse event (binary)<br>4. True bacteremia (binary)<br>5. C-diff associated diarrhea (binary)<br>6. Neutropenic fever days (continuous)<br>*Bacterial and non-bacterial | Patients compared according to the following hierarchy, with decreasing level of clinical significance:<br>1. Death+ (time-to-event)<br>2. Cancer relapse, progression, or SMN (time-to-event)<br>3. SIOP grade $\geq 2$ hearing loss (time-to-event)<br>4. Renal or hematologic targeted toxicity (binary)<br>+All-cause |
| <u>Note</u> | Patients have the best possible outcome if they do not experience an outcome listed over the relevant follow-up time and rank better than those who have any listed outcome in a given pairwise comparison. |  |

### ACCL0934 Hypothetical HCE

For ACCL0934, the first HCE (HCE1) included the following components, ordered from most to least clinically important: 1) Death (all cause), 2) severe infection, 3) BSI, and 4) duration of FN in days. The second HCE (HCE2) was broader, to include potential harms of levofloxacin prophylaxis per the following ranking: 1) Death, 2) severe infection, 3) other serious adverse event (required expedited reporting per-protocol), 4) BSI, 5) *C difficile* diarrhea, and 6) duration of FN. Death specifically attributable to BSI was substituted for all-cause death for analyses presented in the supplement. Days of FN was continuous; all other outcomes were binary.

### ACCL0431 Hypothetical HCE

For ACCL0431, HCE1 pertained to disease and hearing loss outcomes per the following ordering from most to least clinically important: 1) Death, 2) oncologic event (cancer relapse, progression, or a secondary malignancy), and 3) hearing loss. All outcomes were analyzed as time to event. For HCE2 a fourth component was included: 4) one or more of the targeted renal or hematologic toxicities (binary outcome).

Hearing loss was categorized using the International Society of Paediatric Oncology (SIOP) severity grading system as used in post-hoc analyses; this outcome was dichotomized with SIOP Grade ≥ 2 (communication-impacting hearing loss) defined as hearing loss present.^21^ Additional analyses of the HCE were implemented (presented in the supplement), where we instead used the original American Speech-Language-Hearing Association (ASHA) primary outcome.

### Cohorts and Follow-Up Time

For consistency with the original primary manuscripts published for ACCL0934 and ACCL0431, the data definitions and primary analysis datasets were used to the extent possible.

For ACCL0934, analyses were performed separately for acute leukemia and HCT cohorts, and all eligible patients deemed analyzable for the BSI primary outcome in the original publication were included in our analyses. The ACCL0934 primary manuscript did not report on all-cause death, and as such, the primary analysis file was augmented to include this additional outcome for our analyses.^2^ Patients were followed for all HCE component outcomes for the protocol defined infection observation period.

For reanalysis of ACCL0431, all eligible participants except those who withdrew within one day of enrollment were included. Use of SIOP graded hearing loss facilitated inclusion of a larger proportion of the 125 eligible participants with evaluable audiometry (121 with SIOP evaluable audiology as opposed to 104 under the original ASHA defined outcome). Four participants without SIOP gradable hearing evaluations were included but were assumed not to have had a hearing loss event. This ensured participation for analysis of disease outcomes. For the win-statistical analysis of HCE using the non-parametric estimation approach, a fixed follow-up period must be defined. We performed two separate analyses within each of two cohorts: one analysis truncated follow-up at 1-year post-enrollment and the other at 3-years. We chose 1-year to approximate the observation time intended for the original primary ototoxicity outcome. We then sought to evaluate the impact of extended disease outcome monitoring, and we chose 3-years for the purposes of illustration. One cohort included the overall sample of patients, regardless of diagnosis or extent of disease at baseline, which reflects the broader overall target population of the original trial. The second cohort was restricted to patients enrolled with localized disease only, the population for which STS has regulatory approval.^22^

### Statistical Analysis

Descriptive statistics were used to summarize HCE component outcome distributions. Our primary analyses report the win-odds ratio, estimated using the non-parametric patient-level pairwise comparison approach described above. We present estimates of alternative win-statistic metrics in the supplemental materials. For ACCL0934, all patients were considered to have completed the infection observation period if an HCE relevant event was not observed. For ACCL0431, to follow the non-parametric approach, each pairwise comparison utilized the minimum follow-up time between the two patients. Closed form, asymptotic variance estimators were used to compute standard errors, perform hypothesis tests and construct 95% confidence intervals (CI).^16,18^

Visual aids were also produced. For ACCL0934, we utilized a combined bar and forest plot. The bar plot shows the percentage of pairwise comparisons resulting in a win for each arm within each HCE component and the forest plot displays corresponding win-odds effect sizes.^7^ For the ACCL0431 overall cohort analysis, we utilized the Maraca plot as an illustrative visual aid.^23^

All analyses were implemented using R version 4.5.0, employing the WINS and MARACA packages.^24,25^ Additional detail is described and analyses presented in the supplement.

## Results

### Example 1: ACCL0934

The trial enrolled 624 participants; 11 were excluded from analysis due to ineligibility or withdrawal prior to observation start. There were 549 (90%) who completed the entire infection observation period, with the remainder completing between 10-92 days (median 22).

Table 2 summarizes HCE outcome components by arm. In the acute leukemia cohort, 43% of patients randomized to control experienced BSI, whereas 22% experienced BSI on the levofloxacin arm. In the HCT cohort, the BSI cumulative incidences were 17% on the control arm and 11% on the levofloxacin arm. Across HCE component outcomes evaluated, there was a reduction in incidence or duration, whichever relevant, on the levofloxacin relative to control arm, with just two exceptions: there was an equal or nearly equal proportion of deaths in each arm per cohort, and slightly higher proportion of patients who experienced non-infectious serious AEs on levofloxacin relative to control in the HCT cohort (2% versus 0%, respectively).

**Table 2:** ACCL0934 Outcome Summaries and Win-Statistic Analysis Results.

| Event Summaries for HCE Components |  |  |  |  |
| --- | --- | --- | --- | --- |
|  | Acute Leukemia Group |  | HCT Group |  |
|  | Levofloxacin | Control | Levofloxacin | Control |
| **Overall Patients (N)** | 96 | 99 | 210 | 208 |
| **Death (All Cause)**<br>– n (%)<br>(In HCE 1 and HCE 2) | 6 (6%) | 7 (7%) | 25 (12%) | 25 (12%) |
| **Severe Infection**<br>– n (%)<br>(In HCE 1 and HCE 2) | 5 (5%) | 10 (10%) | 6 (3%) | 8 (4%) |
| **Other Serious Adverse Event**<br>– n (%)<br>(In HCE 2 Only) | 1 (1%) | 3 (3%) | 4 (2%) | 0 (0%) |
| **Bloodstream Infection**<br>– n (%)<br>(In HCE 1 and HCE 2) | 21 (22%) | 43 (43%) | 23 (11%) | 36 (17%) |
| **C-diff Associated Diarrhea**<br>– n (%)<br>(In HCE 2 Only) | 2 (2%) | 7 (7%) | 5 (2%) | 9 (4%) |
| **Days of Febrile Neutropenia**<br>– Median (Quartiles)<br>(In HCE 1 and HCE 2) | 4 (2,9) | 6 (3,12) | 2 (0,5) | 3 (1,6) |
| **Patients with <u>No</u> Above-Listed Events and <u>No</u> FN Episode**<br>– n (%) | 12 (13%) | 9 (9%) | 47 (22%) | 20 (10%) |
| Win-Statistic Analysis Results |  |  |  |  |
|  | Acute Leukemia Group |  | HCT Group |  |
| Endpoint | Win-Odds (Ratio) – Estimate (95% CI) | p-value | Win-Odds (Ratio) – Estimate (95% CI) | p-value |
| **BSI Only** | 1.55 (1.17,2.05) | 0.003 | 1.14 (0.99,1.30) | 0.065 |
| **HCE 1** | 1.74 (1.23,2.46) | 0.002 | 1.28 (1.02,1.60) | 0.031 |
| **HCE 2** | 1.77 (1.25,2.50) | 0.001 | 1.27 (1.01,1.58) | 0.037 |

Table 2 also displays estimated win-odds ratios, corresponding 95% CI and p-values. When comparing the BSI endpoint alone across arms using the win statistic approach, randomization to levofloxacin was beneficial (i.e., win odds ratio>1) in both cohorts, but only the ratio for the acute leukemia cohort achieved statistical significance (p-value=0.003). For the analysis of HCE1 the estimated win-odds ratio was in the direction of benefit from levofloxacin relative to control and reached statistical significance in both acute leukemia and HCT cohorts. The effect sizes and p-values under analysis of HCE2 were similar to those of analysis of HCE1 within each cohort, suggesting minimal impact of the added components in HCE2. In an additional analysis where all-cause deaths were replaced with death due to BSI in the HCE analyses, the win-odds effect sizes increased, and corresponding p-values decreased (Supplemental Table 1, HCE1: acute leukemia: 1.83, p-value=0.001; HCT: 1.39, p-value=0.004). The use of alternative win-statistics and methods to calculate standard errors yielded similar conclusions to the results of Table 2 (Supplemental Tables 2 and 3).

Figures 1.1 and 1.2 illustrate the outcomes responsible for the largest percentage of wins across pairwise comparisons within each arm by HCE component. For the acute leukemia cohort, the largest absolute difference in win across arms was observed in the BSI component. In contrast, in the HCT cohort, the largest absolute difference in wins occurred in the FN duration component.

**Figure 1.1.**
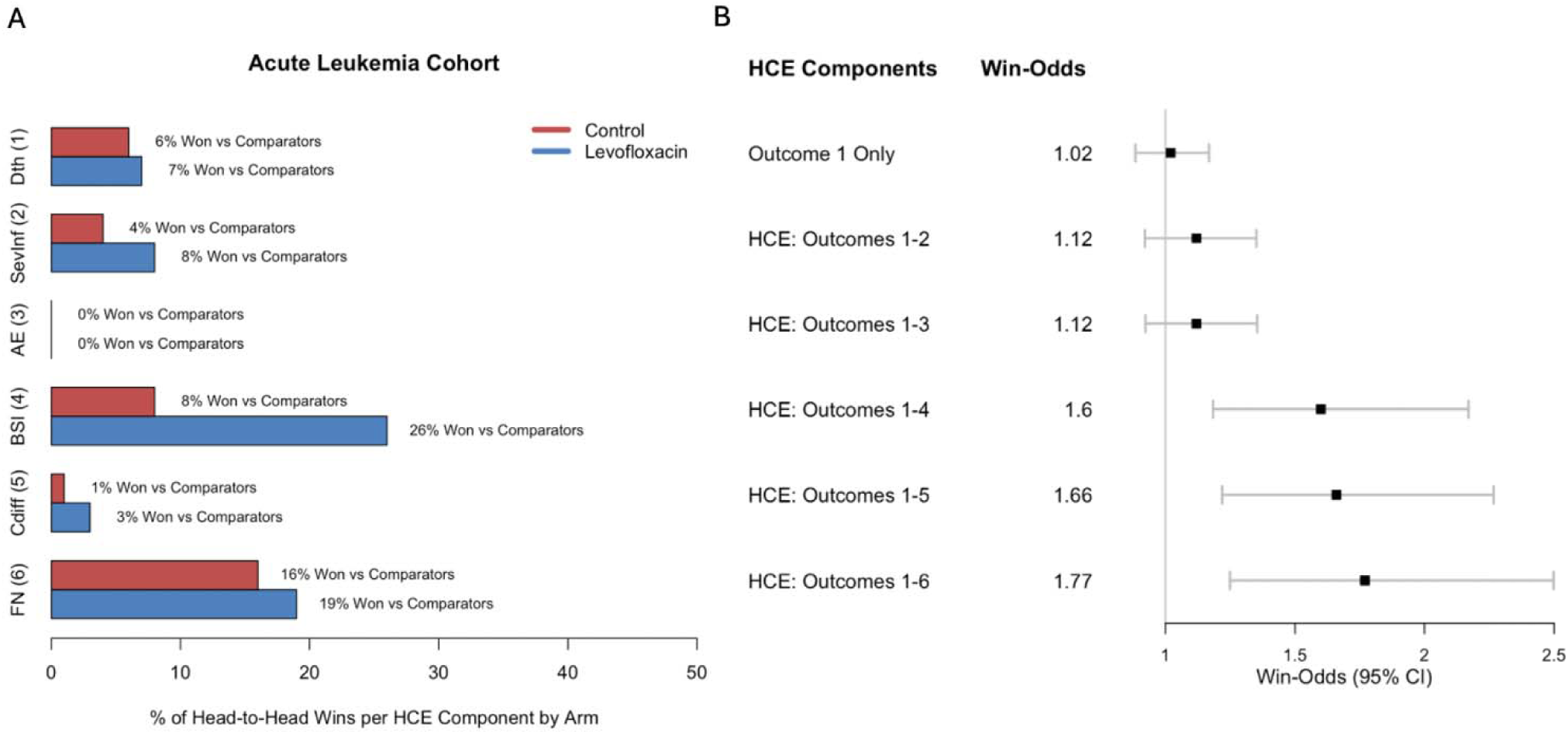
Win percentages and corresponding win-odds ratios in the acute leukemia cohort of ACCL0934. Panel A displays the percentage of patient level pairwise comparisons that resulted in a win within the HCE component displayed, broken up by randomized arm. E.g., across pairwise comparisons performed on the BSI outcome, 26% resulted in a win for the levofloxacin arm patient. Panel B lists the estimated win-odds and plots the point estimate and corresponding 95% confidence interval width in forest plot format. Each win-odds is based on the HCE that includes the outcome listed in the corresponding row of the bar plot (panel A) and all outcomes displayed above, if relevant. Abbreviations: Dth: Death (All Cause); SevInf: Severe Infection; AE: Other severe (non-severe infection) adverse event; BSI: true blood stream infection; Cdiff: *Clostridioides difficile*–associated diarrhea; FN: febrile neutropenia.

**Figure 1.2.**
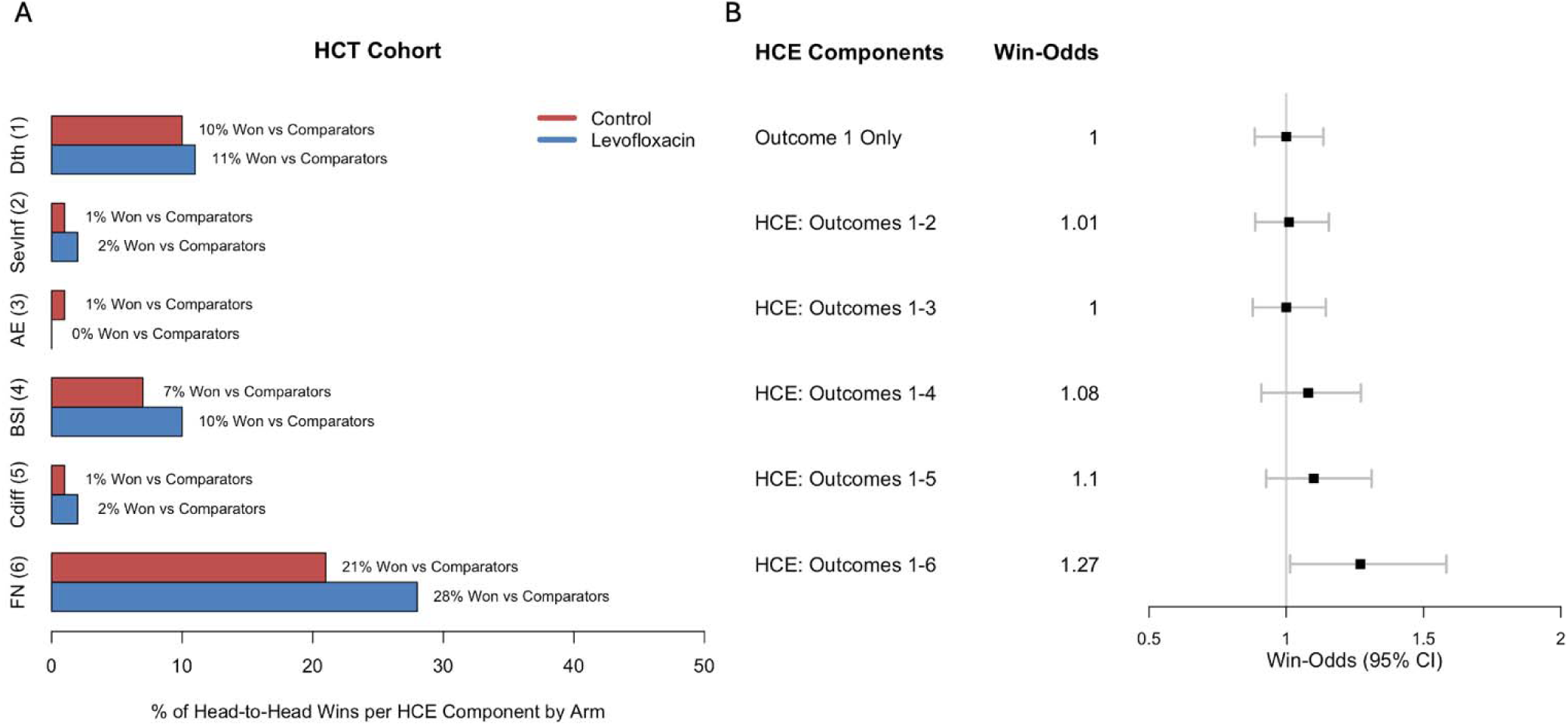
Win percentages and corresponding win-odds in the HCT cohort of ACCL0934. Panel A displays the percentage of patient level pairwise comparisons that resulted in a win within the HCE component displayed, broken up by randomized arm. E.g., across pairwise comparisons performed on the BSI outcome, 10% resulted in a win for the levofloxacin arm patient. Panel B lists the estimated win-odds and plots the point estimate and corresponding 95% confidence interval width in forest plot format. Each win-odds is based on the HCE that includes the outcome listed in the corresponding row of the bar plot (panel A) and all outcomes displayed above, if relevant. Abbreviations: Dth: Death (All Cause); SevInf: Severe Infection; AE: Other severe (non-severe infection) adverse event; BSI: true blood stream infection; Cdiff: *Clostridioides difficile*–associated diarrhea; FN: febrile neutropenia.

### Example 2: ACCL0431

The trial enrolled 125 eligible participants; one was excluded from analyses due study withdrawal on the first day of observation. Among the overall cohort, 4 of the 124 were censored prior to 1-year without a disease or hearing loss event (median follow-up 9-months; range 7-11), and 16 patients were censored earlier than 3-years without experiencing such events (median follow-up 30-months; range 7-35). Among the localized cohort, 3 patients were censored without an event prior to 1-year (median follow-up 9 months; range 7-11), and 11 patients were censored without an event prior to 3-years (median follow-up 34 months; range 7-35).

Table 3 provides descriptive summaries of HCE components. Among the overall cohort, nearly 30% of patients on the control arm experienced clinically significant hearing loss (SIOP grade ≥2), compared to only 5% of patients on the STS arm. The absolute reduction in hearing loss was similar when analyzing the localized disease cohort, where the reduction was from 24% to 5% with follow-up up-to 3-years. Among the overall cohort, there was a larger proportion of patients who died or experienced an oncologic event on the STS arm. This difference was more pronounced when follow-up was extended to 3-years. Regardless of the follow-up duration, there was a similar or identical proportional incidence of death or relapse/disease progression events across arms among the localized disease cohort.

**Table 3:** ACCL0431 Outcome Summaries and Win-Statistic Analysis Results.

| Event Summaries for HCE Components |  |  |  |  |  |  |  |  |
| --- | --- | --- | --- | --- | --- | --- | --- | --- |
|  | Overall Cohort: 1-Year Follow-Up |  | Overall Cohort: 3-Years Follow-Up |  | Localized Cohort: 1-Year Follow-Up |  | Localized Cohort: 3-Years Follow-Up |  |
|  | STS | Control | STS | Control | STS | Control | STS | Control |
| **Overall Patients (N)** | 60 | 64 | 60 | 64 | 38 | 38 | 38 | 38 |
| **Death**<br>– n (%)<br>(In HCE 1 and HCE 2) | 7 (12%) | 6 (9%) | 17 (28%) | 8 (12%) | 3 (8%) | 3 (8%) | 6 (16%) | 4 (11%) |
| **Oncologic Event**<br>– n (%)<br>(In HCE 1 and HCE 2) | 14 (23%) | 12 (19%) | 26 (43%) | 22 (34%) | 8 (21%) | 8 (21%) | 14 (37%) | 13 (34%) |
| **Hearing Loss** – n (%)<br>(In HCE 1 and HCE 2) | 3 (5%) | 17 (27%) | 3 (5%) | 18 (28%) | 2 (5%) | 8 (21%) | 2 (5%) | 9 (24%) |
| **Targeted Renal or Hematologic Toxicity**<br>– n (%)<br>(In HCE 2 Only) | 31 (52%) | 41 (64%) | 31 (52%) | 41 (64%) | 23 (61%) | 29 (76%) | 23 (61%) | 29 (76%) |
| **Patients with No Above-Listed Event**<br>– n (%) | 14 (23%) | 9 (14%) | 7 (12%) | 6 (9%) | 10 (26%) | 5 (13%) | 4 (11%) | 6 (16%) |
| Win-Statistic Analysis Results |  |  |  |  |  |  |  |  |
|  | Overall Cohort: 1-Year Follow-Up |  | Overall Cohort: 3-Years Follow-Up |  | Localized Cohort: 1-Year Follow-Up |  | Localized Cohort: 3-Years Follow-Up |  |
| Endpoint | Win-Odds (Ratio) – Estimate (95% CI) | p-value | Win-Odds (Ratio) – Estimate (95% CI) | p-value | Win-Odds (Ratio) – Estimate (95% CI) | p-value | Win-Odds (Ratio) – Estimate (95% CI) | p-value |
| **Hearing Loss Only** | 1.55<br>(1.17,2.06) | 0.003 | 1.60<br>(1.19,2.15) | 0.002 | 1.38<br>(0.99,1.90) | 0.056 | 1.45<br>(1.03,2.05) | 0.036 |
| **HCE 1** | 1.27<br>(0.89,1.81) | 0.182 | 1.00<br>(0.68,1.45) | 0.985 | 1.20<br>(0.78,1.85) | 0.402 | 1.09<br>(0.69,1.73) | 0.701 |
| **HCE 2** | 1.24<br>(0.84,1.85) | 0.28 | 1.00<br>(0.67,1.49) | 0.994 | 1.06<br>(0.66,1.71) | 0.816 | 1.05<br>(0.65,1.70) | 0.844 |

Oncologic event is defined as cancer relapse, progression, or secondary malignancy. Hearing loss is defined as SIOP grade $\geq 2$ .

The results of the win-odds analyses are also displayed in Table 3. For either duration of follow-up or either cohort analyzed, when hearing loss alone was the endpoint, the estimated win-odds ratios indicated a beneficial effect of randomization to STS versus control (win-odds >1). All corresponding p-values were less than 0.05 except in the 1-year truncated analysis for the localized only cohort (the trial was not powered for this subgroup sample size). For the overall cohort, when follow-up extended to three years, the estimated win-odds ratio was equal to the null (win-odds ratio = 1). For the localized only cohort, the estimated win-odds ratios remained above 1, but the effect sizes differed depending on the follow-up time allowed. The estimated win-odds were similar in the HCE1 and HCE2 analyses, suggesting limited impact of including targeted renal or hematologic toxicities. We considered several alternative analyses for the overall cohort, including use of additional win-statistics, of alternative methods to handle variable censoring times, and analyses using the ASHA defined ototoxicity outcome, which yielded similar conclusions (Supplemental Tables 4, 5, and 6)

Figures 2.1 and 2.2 display time-to-event curves for components of HCE1 under analysis of the overall cohort using Maraca plots. In each plot, the final panel is used to display censoring times. Figure 2.1 illustrates why the win-odds ratio indicated treatment benefit if follow-up was truncated at 1-year due to near-overlapping survival and oncologic event curves, and fewer hearing loss events observed on STS. Figure 2.2 illustrates the mechanism through which the estimated win-odds ratio converged to the null in the HCE2 analysis: the difference between arms in incidence of death and oncologic events increased over time, balancing out the beneficial effect observed for the hearing loss outcome.

**Figure 2.1:**
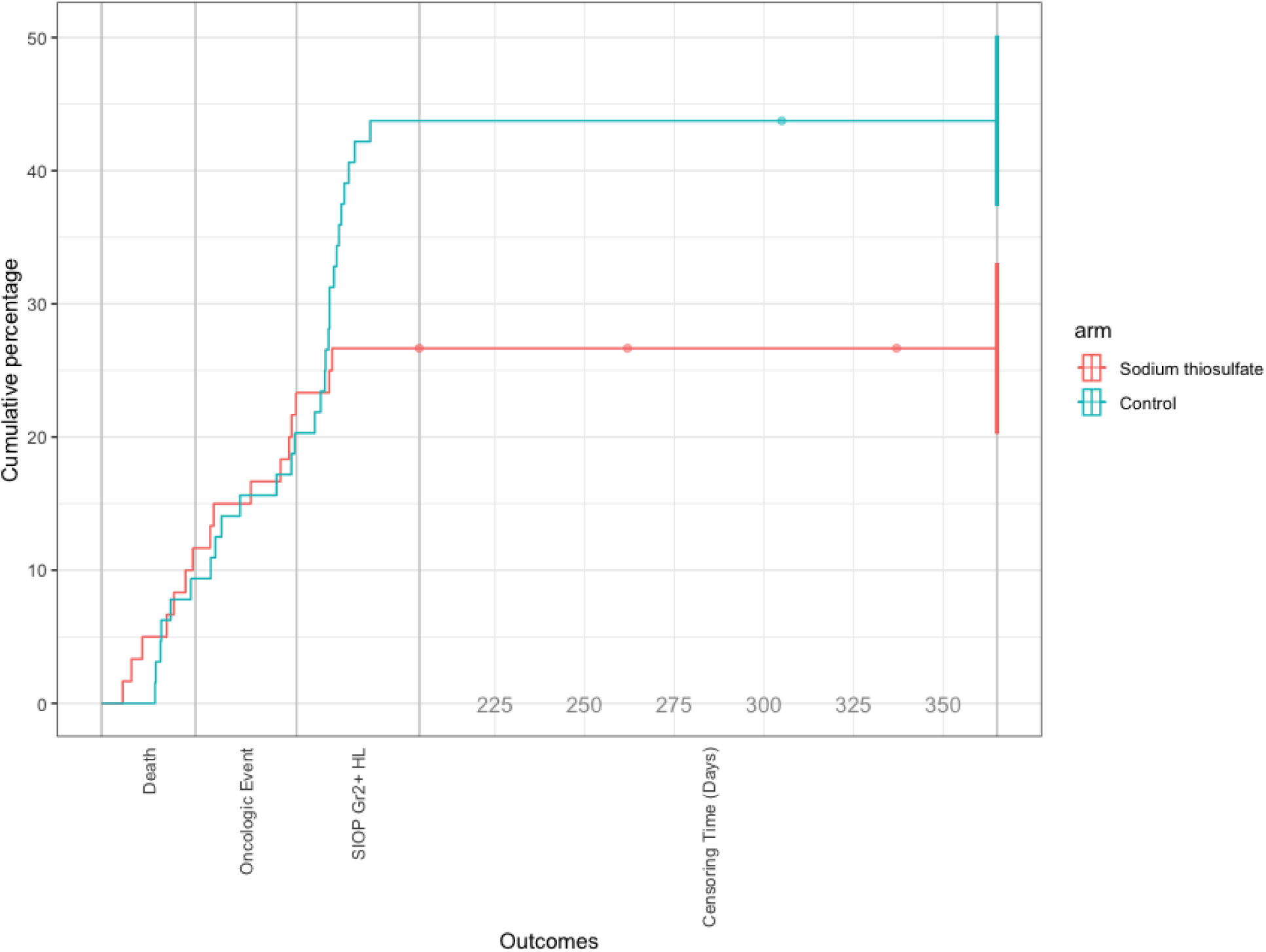
Maraca Plot (Time-to-Event Curves) for HCE Components Among the ACCL0431 Overall Cohort with Follow-Up Truncated at 1-Year The x-axis reflects time, where for each HCE component (death, oncologic event, and hearing loss), time 0 is represented by the bold grey vertical bar on the left of the component, and the maximum follow-up time (1 year) is represented by the bar to the right. For the censoring time column of the figure (right most), the first bold grey vertical bar represents the minimum censoring time observed (approximately 200 days after enrollment). Oncologic events include relapse, secondary malignancy, or disease progression. SIOP Gr2+ HL refers to the binary SIOP grade 2 or higher hearing loss outcome. Censoring Time refers to the time a patient withdrew consent or was last seen if an HCE event hadn’t previously occurred. Except for the time-to-death curve, event curves displayed are among patients who did not experience the events listed to the left.

**Figure 2.2:**
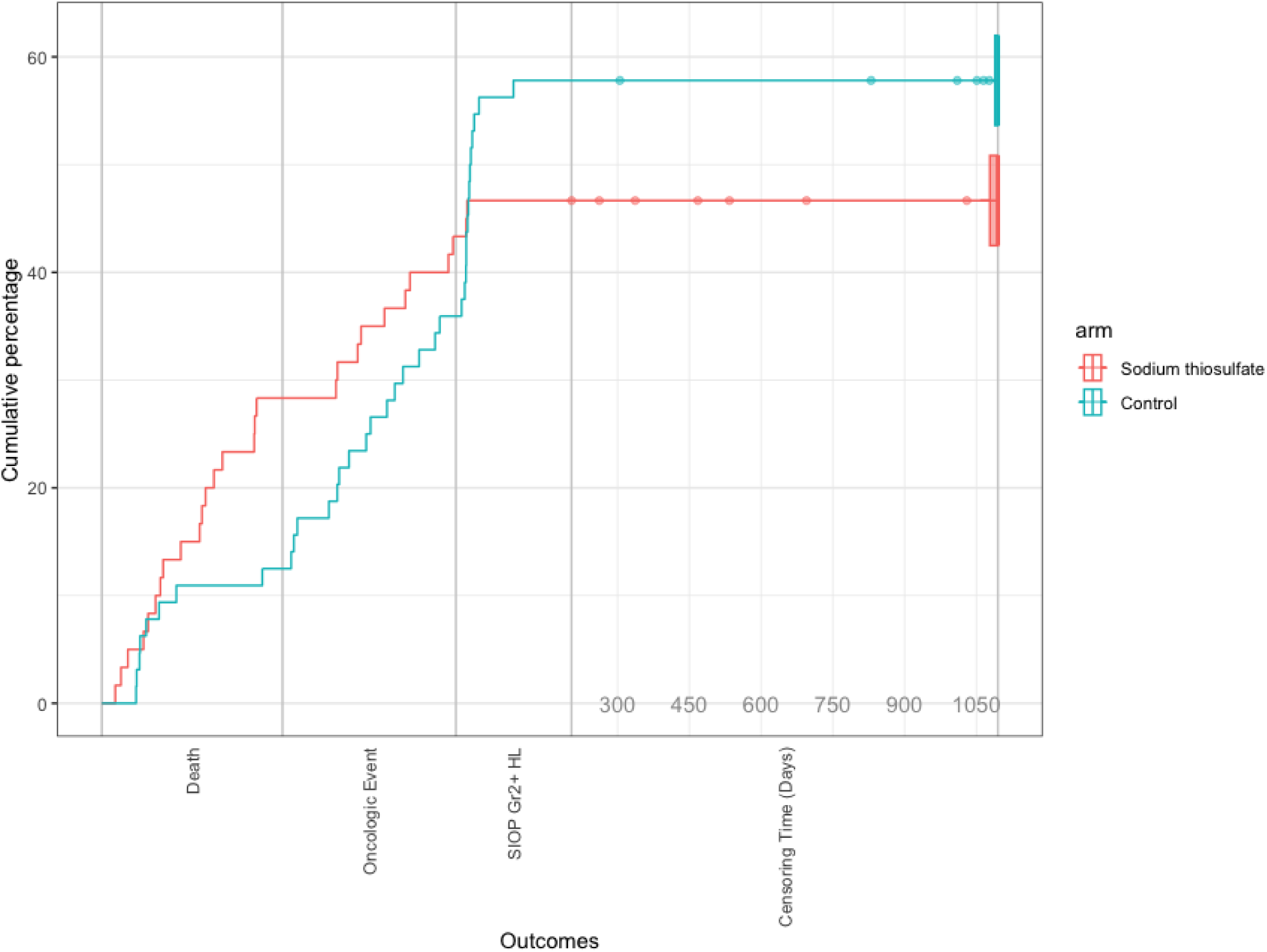
Maraca Plot (Time-to-Event Curves) for HCE Components Among the ACCL0431 Overall Cohort with Follow-Up Truncated at 3-Years. The x-axis reflects time, where for each HCE component (death, oncologic event, and hearing loss), time 0 is represented by the bold grey vertical bar on the left of the component, and the maximum follow-up time (3 years) is represented by the bar to the right. For the censoring time column of the figure (right most), the first bold grey vertical bar represents the minimum censoring time observed (about 200 days after enrollment). Oncologic events include relapse, secondary malignancy, or disease progression. SIOP Gr2+ HL refers to the binary SIOP grade 2 or higher hearing loss outcome. Censoring Time refers to the time a patient withdrew consent or was last seen if an HCE event hadn’t previously occurred. Except for the time-to-death curve, event curves displayed are among patients who did not experience the events listed to the left.

## Discussion

Supportive care trials in pediatric cancer are challenging to design and interpret due to the plurality of key outcomes relevant to interpreting the benefits and harms of randomized treatment.^2,3^ Hierarchical composite endpoints (HCE) and their analysis using win-statistics are powerful but nuanced tools with potential to handle these challenges. These tools are employed to estimate an overall summary measure of the comparative effectiveness of treatments evaluated, incorporating multiple outcomes through a principled approach to handling heterogeneity in clinical importance across those outcomes.^7,13^ We re-analyzed two previously conducted supportive care trials, COG ACCL0934 and ACCL0431, to understand implications of analyses incorporating HCE methods. An overview of lessons learned from our analyses is provided in Table 4.

**Table 4:**
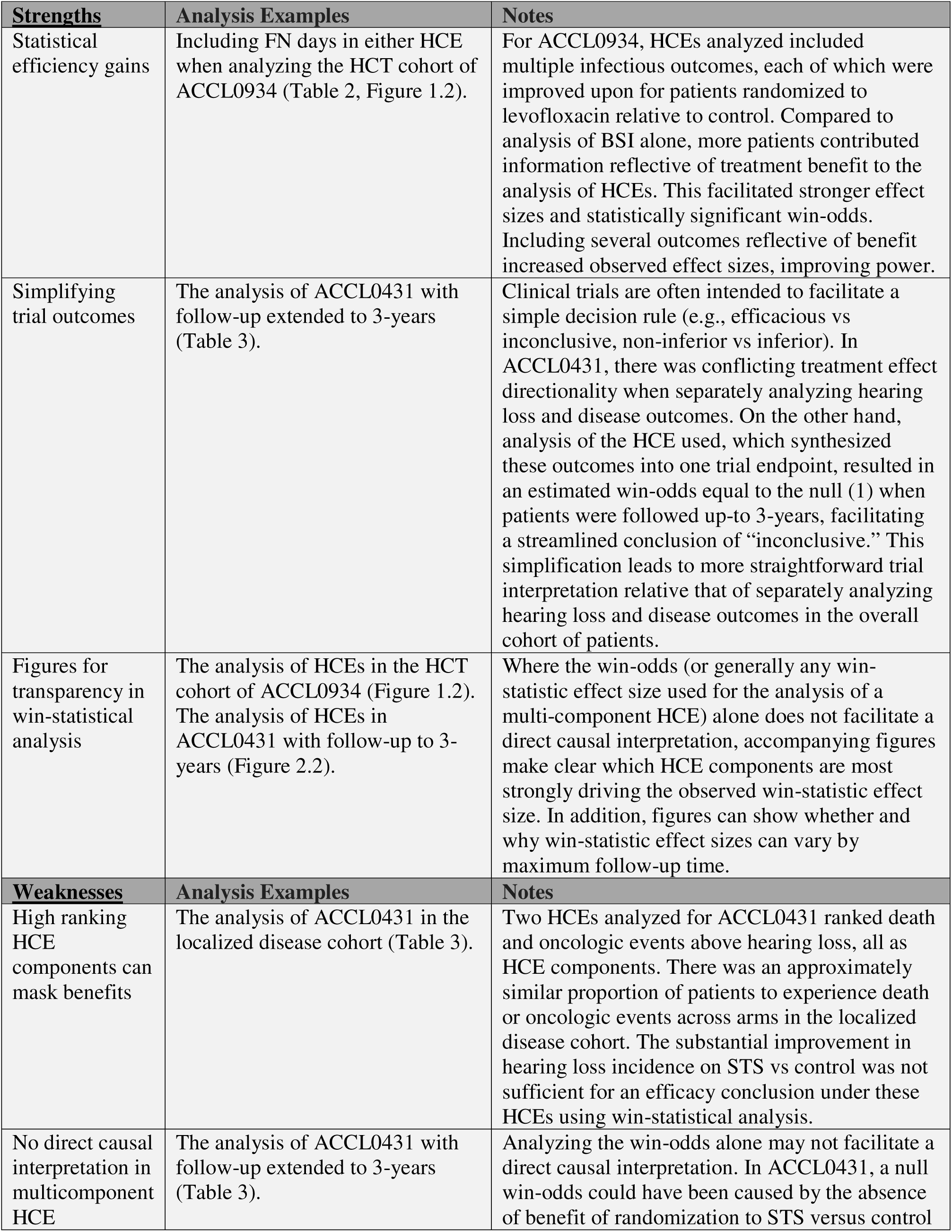
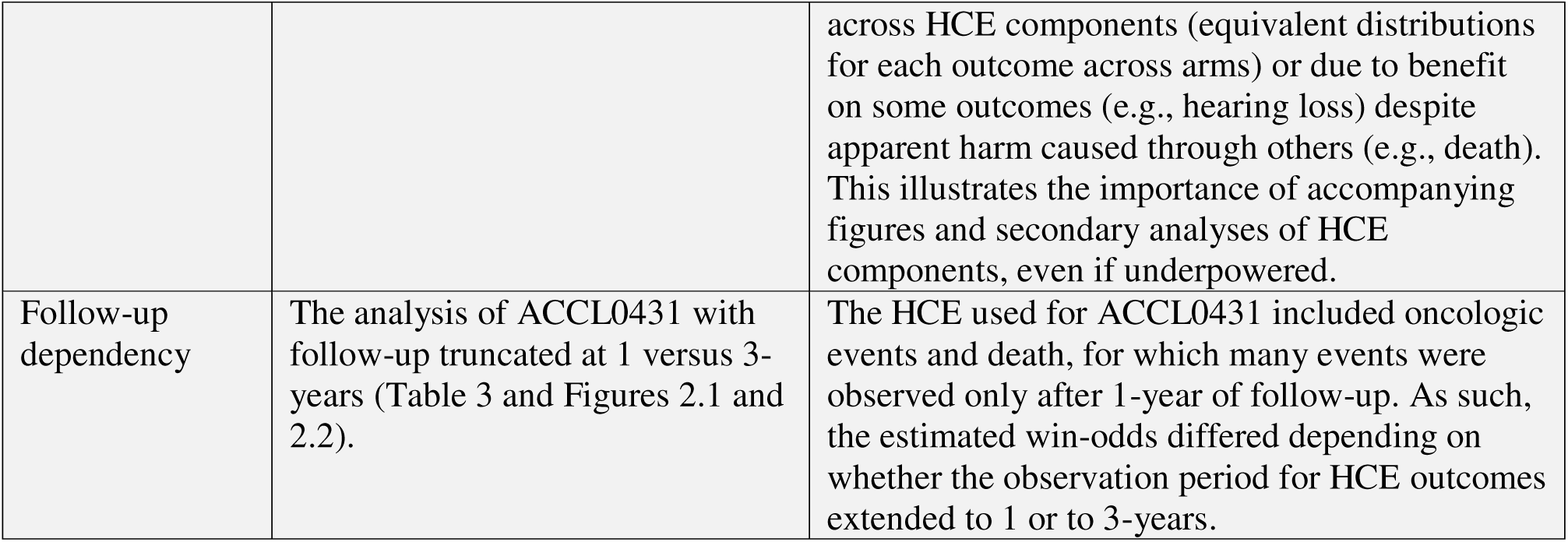
HCE Takeaway Summary.

We emphasize several key findings. Similar to the primary analysis for ACCL0934, when analyzing the BSI primary endpoint alone, the estimated win-odds ratios favored levofloxacin prophylaxis in both cohorts, but the corresponding p-value was only statistically significant (<0.05) for the acute leukemia cohort.^2^ After incorporating additional relevant outcomes into HCEs, the win-odds ratios remained greater than 1 and achieved statistical significance in both cohorts. These analyses of multi-component HCE address a broader clinical question than analysis of BSI alone. The HCEs captured additional benefit of levofloxacin, including reductions of severe infection and durations of time with neutropenic fever. Because each patient contributed more information to the HCE analyses by leveraging additional outcomes, win-statistic effect sizes were larger, yet with sufficiently narrow confidence intervals to allow for statistically significant efficacy conclusions.

In the primary manuscript for ACCL0431, a statistically significant reduction in the proportional incidence of hearing loss was reported for the STS compared to control arm.^3^ On the other hand, considering all participants in aggregate, a trend towards lower survival was estimated on STS versus control. The conflicting directionality in the treatment effect on these outcomes poses challenges to trial interpretation. Our analyses that leverage an HCE inclusive of both ototoxicity and oncologic outcomes in a hierarchical manner facilitate the conclusion that patients from the broader cohort with either localized or disseminated disease randomized to STS do not have higher odds of overall benefit than those randomized to control. This harmonized interpretation in a setting with conflicting directionality of treatment effect on different clinically important outcomes could be viewed as a strength of the HCE approach. This appealing quality of HCE analyses may be generalizable to other supportive care trials which go on to exhibit both benefits and harms of intervention under investigation.

We also conducted analyses in the localized tumor cohort of ACCL0431, which highlighted different considerations. In this cohort, death and oncologic outcomes were similarly distributed across randomized arms, but a larger total number of such events were observed when follow-up was longer. Here, we may expect the win-odds ratios to indicate benefit of randomization to STS due to the ototoxicity reduction and similar distributions of death and oncologic events. Using longer follow-up led to weaker win-odds effect sizes under HCE analyses. This reflects a known property of win-statistical methods. When the highest-ranked HCE outcome occurs more frequently, even if equivalently distributed between arms, the pairwise comparison methodology will find the two groups as more similar, attenuating the estimated effect size. Consequently, a strong intervention effect on lower-ranked outcomes may have limited impact on the win-statistic estimate of the treatment effect on the overall HCE. While clinical relevance must guide HCE construction, statistical efficiency remains an important consideration. Certain HCE structures, such as those evaluated in ACCL0431, may require impractically large sample sizes if the patient populations under consideration are at high-risk for events such as disease relapse.

There are additional important takeaways from our analyses. While win-odds ratios provide a single measure of effect size for the comparison of two study arms with respect to multiple outcomes, these should not be interpreted in the absence of understanding which components of the HCE differ most markedly across arms; visual aids were critical for such transparency. For example, the figure used for ACCL0934 showed FN duration, the lowest ranking HCE component chosen, was needed for an efficacy conclusion using the win-odds ratio in the HCT cohort. In alignment with the HCE literature, when constructing a trial protocol around HCE we advocate that visual aids be declared in the statistical analysis plan.^7,23^ Next, our analyses of ACCL0431 illustrated the impact of follow-up time. Consider the overall cohort analysis. When follow-up time was limited to 1-year, the estimated win-odds ratios were greater than 1, whereas when follow-up was extended to 3-years the estimated win odds ratios were reduced to the null (1). This illustrates that win-statistic metrics can be follow-up time dependent, which is frequently discussed in the methodologic literature.^15^ Therefore, trials should be designed with an *a priori* defined, strongly justified follow-up period, wherein it is expected that most patients enrolled will either reach HCE relevant events or the end of follow-up.

We also consider our analyses in the context of challenges emphasized in the HCE literature. There is an abundance of work describing the role of censoring in win-statistical analysis.^15,26,27^ To motivate this issue, consider that the pairwise comparison estimation procedure cannot be employed in a straightforward way when trial participants do not share a common follow-up. While there are statistical methods proposed for win-statistical inference in these scenarios, they can be more challenging to implement or may introduce barriers in trial design.^15,27^ Conveniently, many supportive care applications give rise to short, well-defined follow-up periods (e.g. monitoring thrombotic events in ALL induction^28^ or infection during neutropenia). ACCL0934, among other infectious outcome trials conducted with COG, serve as important cases in demonstrating the feasibility in following patients for such durations.^2,4,29,30^

Another challenge to the interpretation of analyses of HCE is related to variable risk for HCE components. For example, in trials such as ACCL0431 with a heterogeneous patient population, subgroups may be at different risk for different HCE components (e.g., some disease groups had higher risk for disease relapse or progression). In such settings, a win-statistic effect size can have a fundamentally different interpretation within different subgroups of patients and hence, caution must be applied in pooling across those subgroups. In some cases, trials might include only a patient population believed to have homogeneous risk for HCE components. Alternatively, protocol should specify, *a priori*, intent to be powered for win-statistical subgroup analyses. The methods literature summarizes analytic options.^17,31,32^

Regarding our re-analyses, several limitations must be recognized, which arise from performing post-hoc analyses. Firstly, we recognize that neither trial was designed for an HCE analysis. The implications of our analyses are dependent upon trial designs not intended for the use of HCE. Second, the study team had knowledge of the relevant outcome distributions, by arm, prior to constructing the HCEs. Third, prevailing limitations in the data could not be overcome. For example, additional outcomes may have been considered for the HCEs if not for the absence of relevant data, such as other (non-BSI, non-severe) bacterial infections in ACCL0934. For ACCL0431, an additional limitation, which HCE analysis cannot overcome, is the possibility of unbalanced randomization in prognostic factors as a potential explanation for observed survival differences across STS and control groups with disseminated disease, as discussed extensively.^33^ Considering the many outcomes collected under each trial analyzed, alternative HCE construction approaches could have been considered. The HCE used for this paper illustrate several compelling examples to motivate continued investigation into this approach. An exhaustive evaluation was beyond our intended scope.

## Conclusion

This paper is intended to serve as a useful starting point towards prospective implementation of HCE in pediatric cancer supportive care clinical trials, when appropriate. HCE can be constructed to capture the potential multidimensional benefits and harms of investigational supportive care treatments. The primary strength of the corresponding win-statistical analysis is in harmonizing trial conclusions under the use of a single summary measure of comparative effectiveness. Importantly, the process of choosing HCE components and in assigning clinical hierarchy may undeniably be challenging. Hierarchical composite outcomes hinge critically on both the selection of components and the order in which they are prioritized, as these choices shape the interpretation and perceived significance of results. If stakeholders contest the ranking or relevance of components, the usefulness of conclusions may be questionable. Ultimately, the robustness of such analyses depends on consensus around what matters most. Notably, every clinical trial protocol ranks outcomes by assigning outcomes to primary, secondary, and exploratory aims. Constructing HCE is one of many potential opportunities for a principled approach to the structuring of outcomes inherent to pediatric supportive care clinical trials.

## Disclaimer

The content is solely the responsibility of the authors and does not necessarily represent the official views of the National Institutes of Health.

## Data Sharing Statement

The Children’s Oncology Group Data Sharing policy describes the release and use of COG individual subject data for use in research projects in accordance with National Clinical Trials Network (NCTN) Program and NCI Community Oncology Research Program (NCORP) Guidelines. Only data expressly released from the oversight of the relevant COG Data and Safety Monitoring Committee (DSMC) are available to be shared. Data sharing will ordinarily be considered only after the primary study manuscript is accepted for publication. For phase 3 studies, individual- level de-identified datasets that would be su□cient to reproduce results provided in a publication containing the primary study analysis can be requested from the NCTN/NCORP Data Archive at https://nctn-data-archive.nci.nih.gov/. Data are available to researchers who wish to analyze the data in secondary studies to enhance the public health benefit of the original work and agree to the terms and conditions of use. For non-phase 3 studies, data are available following the primary publication. An individual-level de-identified dataset containing the variables analyzed in the primary results paper can be expected to be available upon request. Requests for access to COG protocol research data should be sent to. Data are available to researchers whose proposed analysis is found by COG to be feasible and of scientific merit and who agree to the terms and conditions of use. For all requests, no other study documents, including the protocol, will be made available and no end date exists for requests. In addition to above, release of data collected in a clinical trial conducted under a binding collaborative agreement between COG or the NCI Cancer Therapy Evaluation Program (CTEP) and a pharmaceutical/biotechnology company must comply with the data sharing terms of the binding collaborative/contractual agreement and must receive the proper approvals.

## References

1. Esbenshade AJ, Sung L, Brackett J, et al. Children’s Oncology Group’s 2023 blueprint for research: Cancer control and supportive care. Pediatr Blood Cancer. 2023;70 Suppl 6(Suppl 6):e30568. doi:10.1002/pbc.30568

2. Alexander S, Fisher BT, Gaur AH, et al. Effect of Levofloxacin Prophylaxis on Bacteremia in Children With Acute Leukemia or Undergoing Hematopoietic Stem Cell Transplantation: A Randomized Clinical Trial. JAMA. 2018;320(10):995–1004. doi:10.1001/jama.2018.12512

3. Freyer DR, Chen L, Krailo MD, et al. Effects of sodium thiosulfate versus observation on development of cisplatin-induced hearing loss in children with cancer (ACCL0431): a multicentre, randomised, controlled, open-label, phase 3 trial. Lancet Oncol. 2017;18(1):63–74. doi:10.1016/S1470-2045(16)30625-8

4. Fisher BT, Zaoutis T, Dvorak CC, et al. Effect of Caspofungin vs Fluconazole Prophylaxis on Invasive Fungal Disease Among Children and Young Adults With Acute Myeloid Leukemia. JAMA. 2019;322(17):1673–1681. doi:10.1001/jama.2019.15702

5. Freyer DR, Orgel E, Knight K, Krailo M. Special considerations in the design and implementation of pediatric otoprotection trials. J Cancer Surviv. 2023;17(1):4–16. doi:10.1007/s11764-022-01312-x.

6. Pocock SJ, Ariti CA, Collier TJ, Wang D. The win ratio: a new approach to the analysis of composite endpoints in clinical trials based on clinical priorities. Eur Heart J. 2012;33(2):176–182. doi:10.1093/eurheartj/ehr352

7. Pocock SJ, Gregson J, Collier TJ, Ferreira JP, Stone GW. The win ratio in cardiology trials: lessons learnt, new developments, and wise future use. Eur Heart J. 2024;45(44):4684–4699. doi:10.1093/eurheartj/ehae647

8. Walker H, McLeman L, Meyran D, et al. Co-designing a Novel Ordinal Endpoint for an Adaptive Platform Trial, BANDICOOT, in Pediatric Hematopoietic Stem Cell Transplant. Transplant Cell Ther. 2025;31(5):321.e1–321.e12. doi:10.1016/j.jtct.2025.01.894

9. Maurer MS, Schwartz JH, Gundapaneni B, et al. Tafamidis Treatment for Patients with Transthyretin Amyloid Cardiomyopathy. N Engl J Med. 2018;379(11):1007–1016. doi:10.1056/NEJMoa1805689

10. Kosiborod MN, Esterline R, Furtado RHM, et al. Dapagliflozin in patients with cardiometabolic risk factors hospitalised with COVID-19 (DARE-19): a randomised, double-blind, placebo-controlled, phase 3 trial. Lancet Diabetes Endocrinol. 2021;9(9):586–594. doi:10.1016/S2213-8587(21)00180-7

11. Gillmore JD, Judge DP, Cappelli F, et al. Efficacy and Safety of Acoramidis in Transthyretin Amyloid Cardiomyopathy. N Engl J Med. 2024;390(2):132–142. doi:10.1056/NEJMoa2305434

12. Sorajja P, Whisenant B, Hamid N, et al. Transcatheter Repair for Patients with Tricuspid Regurgitation. N Engl J Med. 2023;388(20):1833–1842. doi:10.1056/NEJMoa2300525

13. Gasparyan SB, Buenconsejo J, Kowalewski EK, et al. Design and Analysis of Studies Based on Hierarchical Composite Endpoints: Insights from the DARE-19 Trial. Ther Innov Regul Sci. 2022;56(5):785–794. doi:10.1007/s43441-022-00420-1

14. Evans SR, Rubin D, Follmann D, et al. Desirability of Outcome Ranking (DOOR) and Response Adjusted for Duration of Antibiotic Risk (RADAR). Clin Infect Dis Off Publ Infect Dis Soc Am. 2015;61(5):800–806. doi:10.1093/cid/civ495

15. Mao L. Defining estimand for the win ratio: Separate the true effect from censoring. Clin Trials Lond Engl. 2024;21(5):584–594. doi:10.1177/17407745241259356

16. Dong G, Li D, Ballerstedt S, Vandemeulebroecke M. A generalized analytic solution to the win ratio to analyze a composite endpoint considering the clinical importance order among components. Pharm Stat. 2016;15(5):430–437. doi:10.1002/pst.1763

17. Gasparyan SB, Folkvaljon F, Bengtsson O, Buenconsejo J, Koch GG. Adjusted win ratio with stratification: Calculation methods and interpretation. Stat Methods Med Res. 2021;30(2):580–611. doi:10.1177/0962280220942558

18. Bebu I, Lachin JM. Large sample inference for a win ratio analysis of a composite outcome based on prioritized components. Biostat Oxf Engl. 2016;17(1):178–187. doi:10.1093/biostatistics/kxv032

19. Dong G, Huang B, Verbeeck J, et al. Win statistics (win ratio, win odds, and net benefit) can complement one another to show the strength of the treatment effect on time-to-event outcomes. Pharm Stat. 2023;22(1):20–33. doi:10.1002/pst.2251

20. Brunner E, Vandemeulebroecke M, Mütze T. Win odds: An adaptation of the win ratio to include ties. Stat Med. 2021;40(14):3367–3384. doi:10.1002/sim.8967

21. Orgel E, Knight KR, Villaluna D, et al. Reevaluation of sodium thiosulfate otoprotection using the consensus International Society of Paediatric Oncology Ototoxicity Scale: A report from the Children’s Oncology Group study ACCL0431. Pediatr Blood Cancer. Published online July 7, 2023:e30550. doi:10.1002/pbc.30550

22. FDA Approves Sodium Thiosulfate to Reduce the Risk of Ototoxicity Associated with Cisplatin in Pediatric Patients with Localized, Non-Metastatic Solid Tumors. U.S. Food and Drug Administration.; 2024. https://www.fda.gov/drugs/resources-information-approved-drugs/fda-approves-sodium-thiosulfate-reduce-risk-ototoxicity-associated-cisplatin-pediatric-patients?utm_source=chatgpt.com

23. Karpefors M, Lindholm D, Gasparyan SB. The maraca plot: A novel visualization of hierarchical composite endpoints. Clin Trials Lond Engl. 2023;20(1):84–88. doi:10.1177/17407745221134949

24. Cui Y, Huang B. WINS: The R WINS Package.

25. Karpefors M, Gasparyan SB, Borini S, Huhn M. maraca: The Maraca Plot: Visualizing Hierarchical Composite Endpoints. Published online June 8, 2025.

26. Li H, Chen WC, Lu N, Tang R, Zhao Y. The Elusiveness of the Win Ratio Parameter in the Presence of Missing Data. Ther Innov Regul Sci. 2024;58(3):431–432. doi:10.1007/s43441-024-00645-2

27. Dong G, Mao L, Huang B, et al. The inverse-probability-of-censoring weighting (IPCW) adjusted win ratio statistic: an unbiased estimator in the presence of independent censoring. J Biopharm Stat. 2020;30(5):882–899. doi:10.1080/10543406.2020.1757692

28. O’Brien SH, Rodriguez V, Lew G, et al. Apixaban versus no anticoagulation for the prevention of venous thromboembolism in children with newly diagnosed acute lymphoblastic leukaemia or lymphoma (PREVAPIX-ALL): a phase 3, open-label, randomised, controlled trial. Lancet Haematol. 2024;11(1):e27–e37. doi:10.1016/S2352-3026(23)00314-9

29. Zerr DM, Milstone AM, Dvorak CC, et al. Chlorhexidine gluconate bathing in children with cancer or those undergoing hematopoietic stem cell transplantation: A double blinded randomized controlled trial from the Children’s Oncology Group. Cancer. 2021;127(1):56–66. doi:10.1002/cncr.33271

30. Dvorak CC, Fisher BT, Esbenshade AJ, et al. A Randomized Trial of Caspofungin vs Triazoles Prophylaxis for Invasive Fungal Disease in Pediatric Allogeneic Hematopoietic Cell Transplant. J Pediatr Infect Dis Soc. 2020;10(4):417–425. doi:10.1093/jpids/piaa119

31. Song J, Verbeeck J, Huang B, et al. The win odds: statistical inference and regression. J Biopharm Stat. 2023;33(2):140–150. doi:10.1080/10543406.2022.2089156

32. Wang D, Zheng S, Cui Y, He N, Chen T, Huang B. Adjusted win ratio using the inverse probability of treatment weighting. J Biopharm Stat. 2025;35(1):21–36. doi:10.1080/10543406.2023.2275759

33. Orgel E, Villaluna D, Krailo MD, Esbenshade A, Sung L, Freyer DR. Sodium thiosulfate for prevention of cisplatin-induced hearing loss: updated survival from ACCL0431. Lancet Oncol. 2022;23(5):570–572. doi:10.1016/S1470-2045(22)00155-3

